# Diagnosing Left Bundle Branch Block in 12-lead Electrocardiogram using Self-Attention Convolutional Neural Networks

**DOI:** 10.1101/2023.06.25.23291867

**Authors:** Alireza Sadeghi, Alireza Rezaee, Farshid Hajati

## Abstract

The left bundle branch block is a cardiac conduction disorder that affects the heart’s electrical system. It causes the left ventricle of the heart to contract later than the right ventricle, leading to an irregular heartbeat. The diagnosis of left bundle branch block is crucial in determining the most effective treatment for heart irregularities, including cardiac resynchronization therapy. Cardiac resynchronization therapy uses a pacemaker-like device to resynchronize the heart’s contractions and improve its function. However, diagnosing left bundle branch block accurately can be challenging using traditional diagnostic methods that rely on electrocardiograms. This study introduces Self-Attention Convolutional Neural Networks for the detection of left bundle branch block from 12-lead electrocardiograms data using SE-Residual blocks and a self-attention mechanism to highlight important input data features for more accurate diagnosis of left bundle branch block. The model is trained and validated on a database of 10,344 12-lead electrocardiogram samples using a 10-fold cross-validation approach. The results demonstrate an accuracy of 98.91% ± 0.1%, specificity of 99.28% ± 0.15%, precision of 73.09% ± 3.5%, recall or sensitivity of 82.83% ± 4.34%, F1 score of 77.5% ± 1.59%, and area under the receiver operating characteristics curve of 0.91 ± 0.02. The experimental results demonstrate that the proposed deep learning model achieves high accuracy, specificity, and F1 score. These findings suggest that the proposed model can serve as an effective diagnostic tool for identifying left bundle branch block with a high level of efficiency, improving the diagnostic process, and promoting early treatment in medical settings.

## 1. INTRODUCTION

Cardiac resynchronization therapy (CRT) is a medical procedure that involves the placement of a pacemaker in the body to correct irregular heartbeats [1]. This therapy has been found to improve the health of heart patients and increase their lifespan [2, 3]. It is generally more effective in individuals with a QRS complex length of over 120 milliseconds on their electrocardiogram (ECG). Moreover, research suggests that a wider QRS complex or the presence of left bundle branch block (LBBB) on the ECG affect CRT procedure [3–6]. Therefore, the presence or absence of LBBB is critical in determining the efficacy of CRT in heart disease. LBBB is a heart rhythm disorder that is characterized by faulty left ventricular contraction due to a partial or complete malfunction of the electrical system on the left side of the heart. The American Heart College and the American Heart Association provide specific criteria for identifying LBBB on an ECG [7]. However, numerous studies [3, 5, 8–10] have shown that conventional methods and criteria for diagnosing LBBB are prone to errors and mistakes in over 30% of cases. Hence, an accurate and comprehensive system for ECG data analysis and LBBB diagnosis is crucial.

Computer Aided Diagnosis (CAD), used since 1966, is a field of study that uses advanced technology to assist physicians in diagnosing diseases and its objective is to improve the accuracy and efficiency of medical diagnoses [11, 12]. The use of CAD systems has many benefits, including increased accuracy and efficiency of diagnoses, reduced errors and variability between physicians, and improved patient outcomes [13]. The development of CAD systems also has the potential to reduce healthcare costs by reducing the need for unnecessary tests and procedures [13]. Recently, deep neural networks have become essential components of intelligent CAD systems as they enable computers to learn from data and improve their accuracy over time [14, 15]. Deep learning algorithms can recognize complex patterns in medical data and identify subtle changes that may be missed by human eyes.

## 2. RELATED WORKS

The detection of left bundle branch block (LBBB) in electrocardiogram (ECG) data has become increasingly widespread through the use of machine learning and deep learning techniques. Initially, research in this field focused on utilizing small datasets with few channels or leads. For instance, in [16], researchers aimed to classify ECG records from the MIT-BIH dataset [17] into three categories: normal, right bundle branch block (RBBB), and left bundle branch block (LBBB). The dataset included 48 long ECG signals with leads MLII and V5 from 47 patients, and this study utilized 11 noisy signals. The researchers employed the structural and morphological properties of the ECG signals, including RR intervals and PQ intervals, as features for classification. They utilized a Genetic Algorithm (GA) to reduce the number of features, and the final set of features was then classified using a neural network, which achieved a classification accuracy of 98.71%. In addition, the researchers experimented with Principal Component Analysis (PCA) for feature reduction, but the results were not as good as the GA experiment, achieving an accuracy of 77.66%.

In a different study, the goal of [18] was to classify electrocardiogram (ECG) records into three classes: LBBB, RBBB, and normal signals. The researchers utilized 11 samples from the MIT-BIH dataset [17], consisting of 5 normal, 3 LBBB, and 3 RBBB signals. To identify appropriate features for final classification, they introduced the adaptive bacterial forage optimization (ABFO) algorithm, an enhanced version of the bacterial forage optimization (BFO) algorithm [19, 20]. The ABFO algorithm was utilized to eliminate groups with low searching methods, leading to the identification of appropriate groups with effective search strategies. A total of 20 features were produced by the ABFO algorithm, which were then input into the Levenberg–Marquardt Neural Network [21] for classification. The resulting system achieved accurate classification of ECG signals with an accuracy of 98.74%.

In 2018, another study for classifying ECG records into the three classes of LBBB, RBBB, and normal signals was conducted by [22]. In this study, the same MIT-BIH dataset [17] was used. Signals were first filtered using Pan Tompkins algorithm [23] and then different methods including PCA, Magnitude Squared Coherence (MSC) [24], and wavelet transform (WT) [25] were used on them to extract proper features. According to the study, the features generated by PCA were reliable for identifying LBBB, while those derived from MSC were effective in identifying RBBB. Features extracted by WT were also useful in providing stable time and frequency resolutions in noisy environments. To reduce the total number of features, only statistical properties, such as minimum value and median value, were used for the final classification. The final classification was performed using a Random Forest model consisting of 300 decision trees, resulting in an average accuracy of 98.4%.

In spite of previous studies which used hand crafted features for ECG classification, Yildirim et al. [26] proposed an end-to-end model utilizing deep learning techniques to classify 17 different heart arrhythmias, including LBBB. They utilized the same MIT-BIH dataset [17], but only the MLII lead was considered. The researchers generated 1,000 fragments from ECG records of 45 patients and trained the model on this data for 50 epochs. Input signals were first rescaled to the range of - 1 to 1 and then fed to a 16-layer deep model, consisting of 1D convolutional, batch normalization, max-pooling, and dense layers. To address the issue of overfitting, dropout layers [27] were also incorporated. The model demonstrated 100% accuracy in detecting LBBB complications. However, the model was designed to perform effectively at a segment-level input rather than a patient-level input. Additionally, the size of filters used in the convolutional layers was large (7, 10, 15), which may result in slower performance of the system in real-time applications.

The above-mentioned studies relied on small datasets, and often required expert knowledge for feature extraction. With the advent of larger ECG datasets that include signals from multiple leads (such as 3 leads or 6 leads), research has expanded to include the analysis of multi-lead signals. These multi-lead signals can provide a more comprehensive view of the heart’s electrical activity, offering models additional information during the training process. It is worth mentioning that 12-lead ECG signals contain a significant amount of information, making models trained on these signals highly reliable for diagnosing heart arrhythmias.

Ribeiro et al. [28] conducted a study to classify 12-lead ECG records into six different classes, which includes LBBB. Their proposed model includes convolutional, batch normalization, and dense layers, while residual blocks [29] serve as the primary components. The researchers trained the model for 50 epochs on a dataset comprising over two million samples, produced by the Telehealth Network of Minas Gerais (TNMG) [30]. The labels for these samples were only available in textual format, and the researchers utilized an automated method in the first step to extract these labels for each sample in the dataset. Additionally, 98% of the samples in the dataset were utilized for training and validation, while the remaining 2% was used for testing purposes. The proposed model achieved a F1-score of 100% for detecting LBBB.

In 2022, a study was conducted by Chen et al. [31] to classify 12-lead ECG samples into nine different heart arrhythmias, including Bundle Branch Block (both RBBB and LBBB). The authors proposed a deep learning model that consisted of convolutional, batch normalization, residual blocks, dropout, LSTM, attention, and dense layers. The model was trained for 200 epochs using a dataset introduced by Kaohsiung Medical University Hospital (KMUH) [32], which contained 19,253 different samples. To prepare suitable data for the study, the researchers took images from all ECG reports, removed personal information from each image, and digitized the images to produce 12-lead signals. The final model achieved an average accuracy of 96.02% across all arrhythmias. Furthermore, the researchers evaluated their proposed model on the CPSC-2018 dataset [33, 34], which contains 6,877 12-lead ECG signals, and the model achieved an average accuracy of 94.07%.

Previous efforts to use machine learning to diagnose LBBB have several drawbacks and limitations. Firstly, most of these studies require manual extraction of features from ECG data, a process that requires the involvement of experts in the field and increases the application cost. An end-to-end model that can detect LBBB directly from raw ECG data without human intervention would alleviate this issue. Secondly, these studies often use a limited number of training samples, which makes the resulting model vulnerable to overfitting and unreliable in practical applications. Training a deep learning model on a large ECG sample dataset can overcome this limitation and result in a more trustworthy model. Moreover, proposed models in some studies are computationally expensive and may cause many challenges in real-time applications. Lastly, the performance of these models is limited, and errors in diagnosis can have negative impacts on patients’ health, so a highly accurate and reliable system is necessary.

We propose a Self-Attention Convolutional Neural Network (SACNN) that utilizes SE-Residual blocks and attention mechanism for the classification of raw 12-lead ECG data. The ECG data acquired from the 2020 PhysioNet/Computing in Cardiology Challenge [35] was inputted into the SACNN which consisted of 1D convolutional layers, 4 SE-Residual blocks, and a soft-attention block. The SACNN model was then able to accurately identify cases of LBBB in the dataset with an accuracy of 99%.

## 3. MATERIALS AND METHODS

### 3.1. Data

To develop a model for diagnosing LBBB from 12-lead ECG signals, we used the dataset provided by the 2020 PhysioNet/Computing in Cardiology Challenge [35]. Emory University collected this dataset from 10,344 patients in the southeastern United States and includes two sets of files: record files and annotation files. The record files contain 12-lead ECG signals recorded at a 500 Hz sampling frequency ranging from 5 to 10 seconds in length. The annotation files contain information about patient demographics, diagnosis, medical history, symptoms, and surgical history. We created labels for each sample in the record file using the diagnosis information in the annotation file and used them to train the proposed model.

The dataset consists of ECG signals from both male and female patients. It contains 5,551 samples of males (53.66%) and 4,793 samples of females (46.33%). The dataset also exhibits age diversity, with ECG data ranging from 14 to 89 years of age, and the average age of patients is approximately 60 years old. Out of the total number of samples, 231 (2.23%) are found to have LBBB abnormalities in their ECG recordings. Of those 231 samples, 108 (46.75%) belong to male patients, and 123 (53.25%) belong to female patients. Additionally, 81.39% of patients with LBBB are 60 years of age or older. Figure 1 shows the demographic distribution of the studied population.

**Figure 1.**
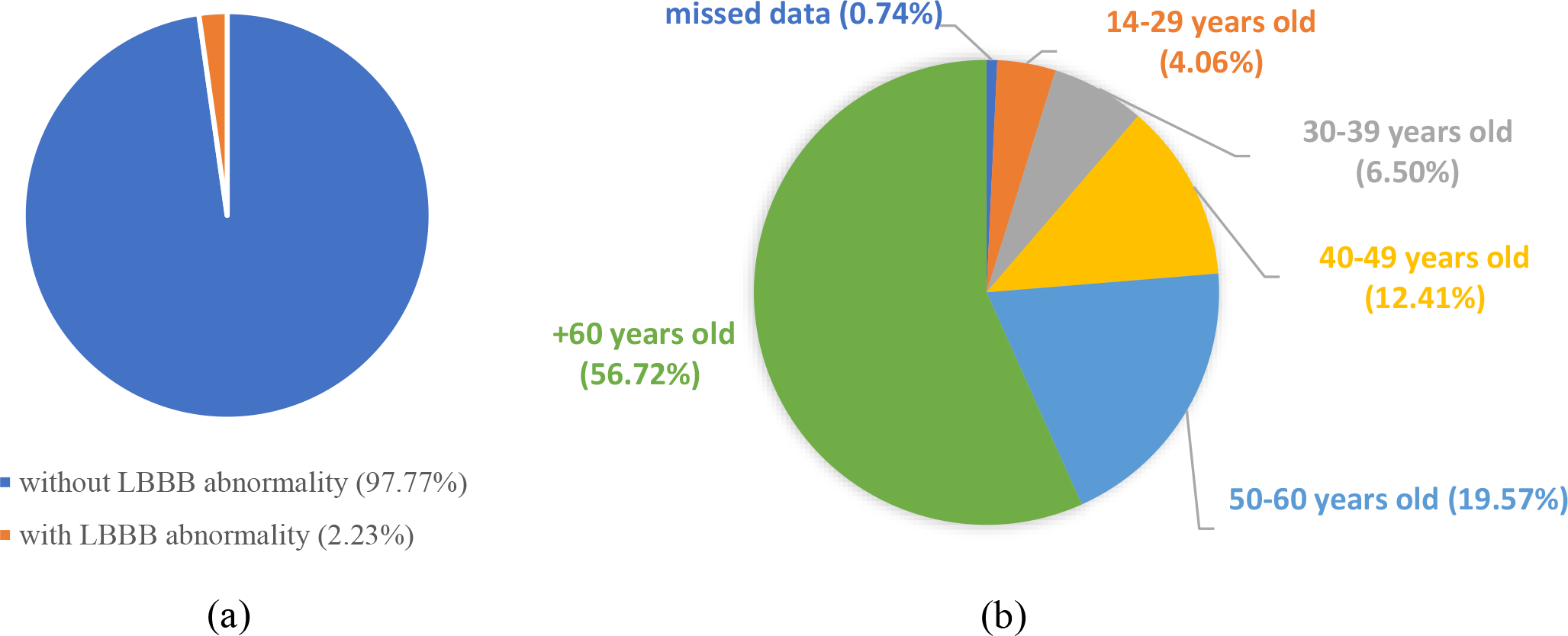
a) LBBB existance ratio, b) Age distribution of patients

We have processed the original data in two steps. First, we normalize the data using the z-score normalization method which addresses the issue of amplitude scaling and offset effect [36]. Second, we standardize the duration of all signals to 8 seconds. Short signals were extended using zero padding, while long ones were truncated to retain just the first 8 seconds (see Figure 2). It should be noted that to enhance the robustness of our model against faulty inputs, we deliberately refrained from removing any compromised ECG signals or utilizing any noise cancellation methods.

**Figure 2.**
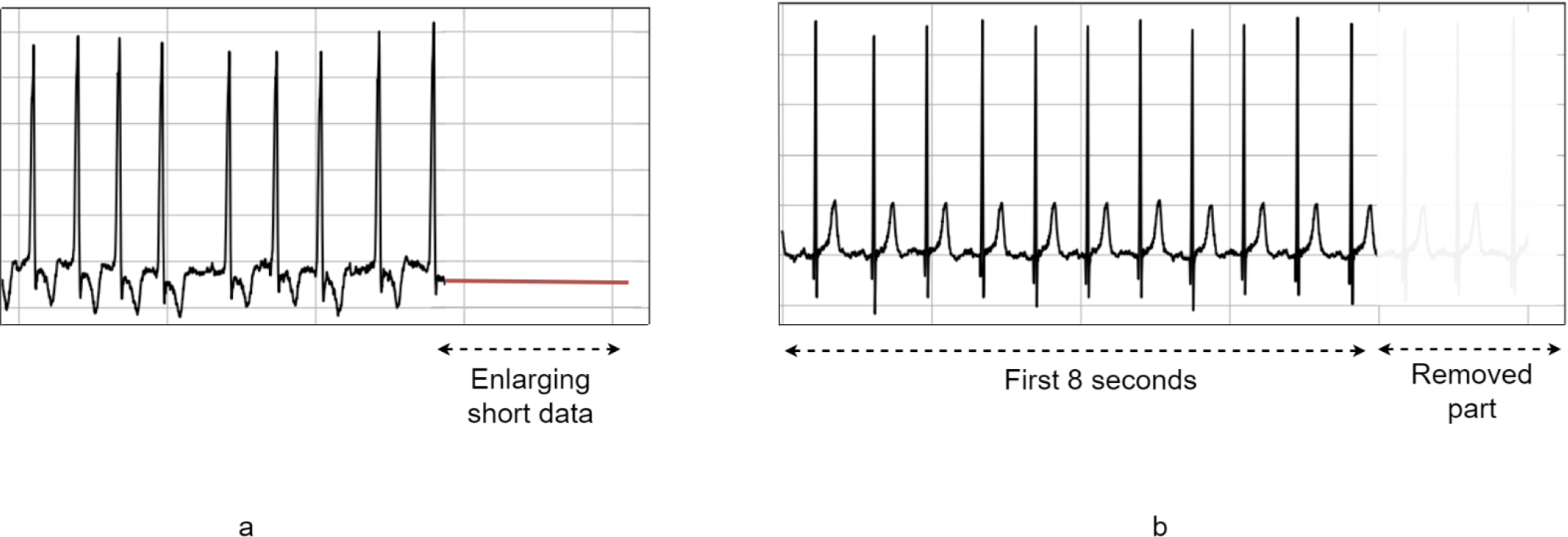
a) Extending short data, b) Truncating long data

### 3.2. Self-Attention Convolutional Neural Network (SACNN)

Figure 3 illustrates the architecture of the proposed soft-attention convolutional neural network (SACNN). In this architecture, several weighted (convolutional) and non-weighted (batch normalization and activation function) layers are first applied to the processed 12-lead ECG signals. Following this, a convolutional layer with a filter size of 3 and stride size of 2 is applied, followed by a pack of 4 SE-Res blocks which are comprehensively discussed later in this section. To reduce the feature size, a max pooling layer with a size of 7 is deployed, after which a self-attention block is utilized. It is worth noting that the filter size of all Conv1D layers utilized in the model is set to 3 which is the smallest size that can effectively capture the information of a single point and its neighborhood. The features extracted through these layers are ultimately fed into a set of fully connected layers for the final classification. Throughout the remainder of this section, we will introduce the various blocks that have been implemented in SACNN in detail.

**Figure 3.**
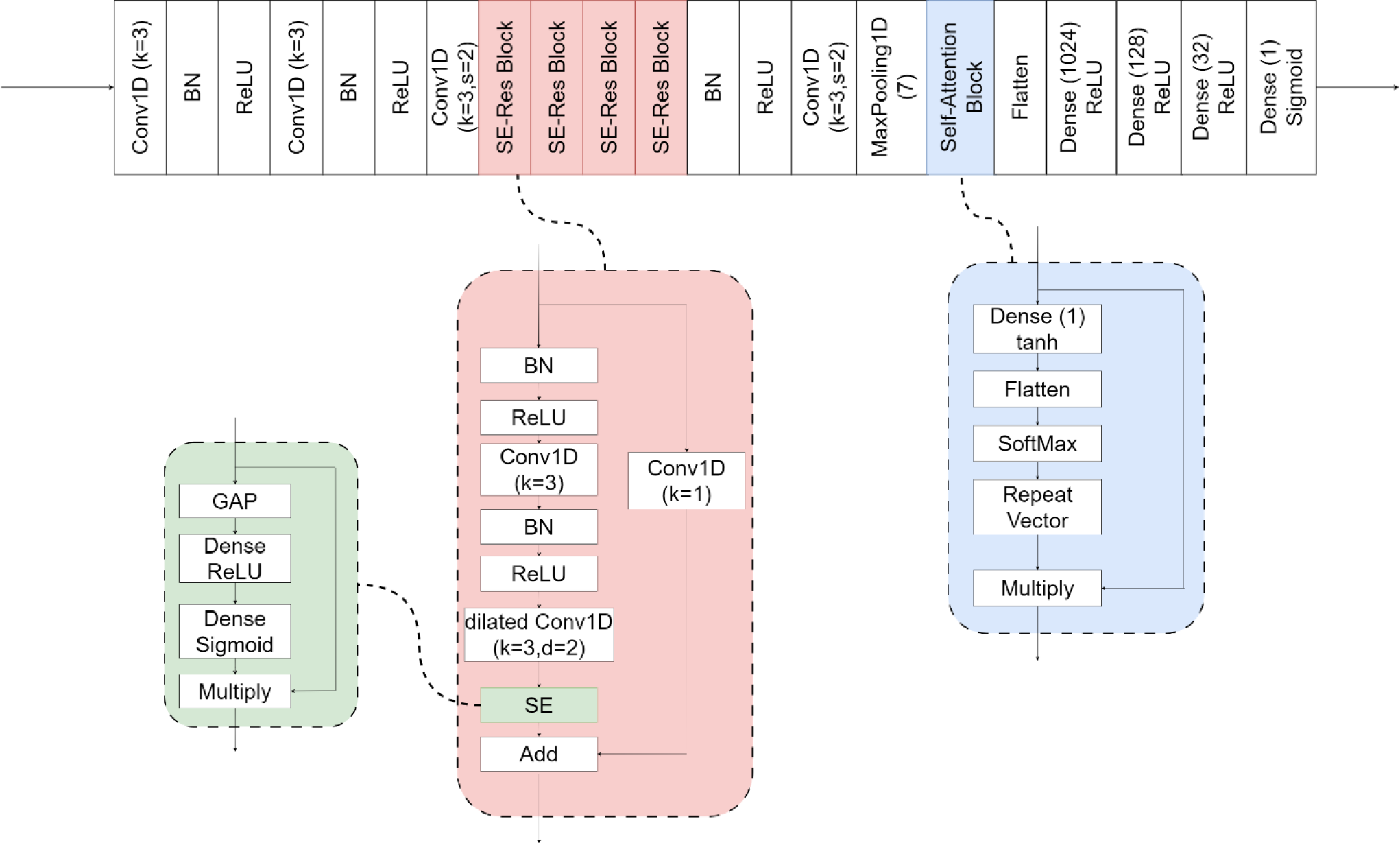
Structure of SACNN (top) which contains several SE-Res blocks (buttom-middle and left) and a self-attention block (buttom-right).

The information contained in different channels of a signal is not equally important. Some channels have a greater impact on the final results than others. To address this challenge, Squeeze and Excitation (SE) blocks have been designed [37]. These blocks magnify the importance of more crucial channels and reduce the importance of less important ones.

The SE blocks consist of two steps. The first step is called squeeze, in which the information of each channel is compressed into a scalar value. The second step is called excitation, which transforms the scalar values obtained in the first step into an array of scalars, denoted as ***s*** that indicate the importance of each channel. Here, Global Average Pooling (GAP) is chosen as the function for the transformation in the first step, and a neural network is used to determine the values of the array ***s***.

Every input signal to a SE block is represented as *X* ∈ *R*^*m*×*n*^, where *n* is the number of channels and *m* is the number of features in each channel.

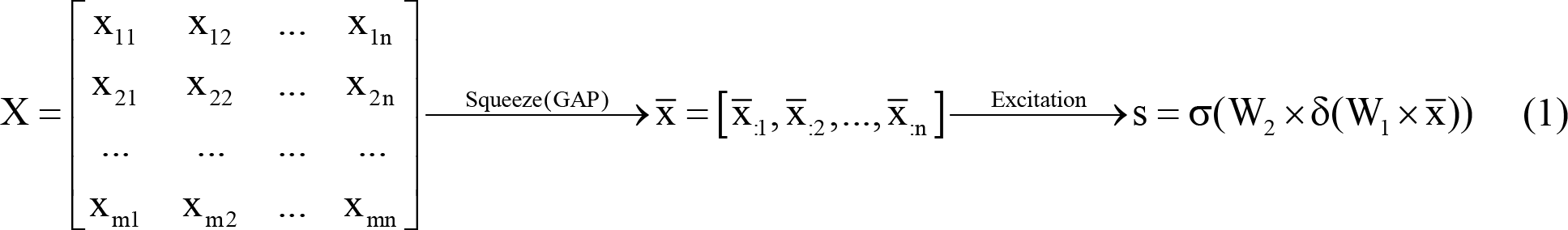

where σ and δ are Sigmoid and ReLU [38] activation functions, respectively. During the training process, the values of 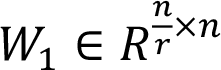 and 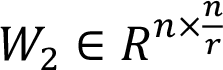 are adjusted to achieve the desired values. In the proposed model, the *r* parameter is set to 8. The array s contains scalar values, each indicating the importance of its corresponding channel. every channel in *X* is multiplied by its corresponding value in the array s and a new tensor denoted as *X*^′^is generated and used as the input for the next layer.

**Figure 4.**
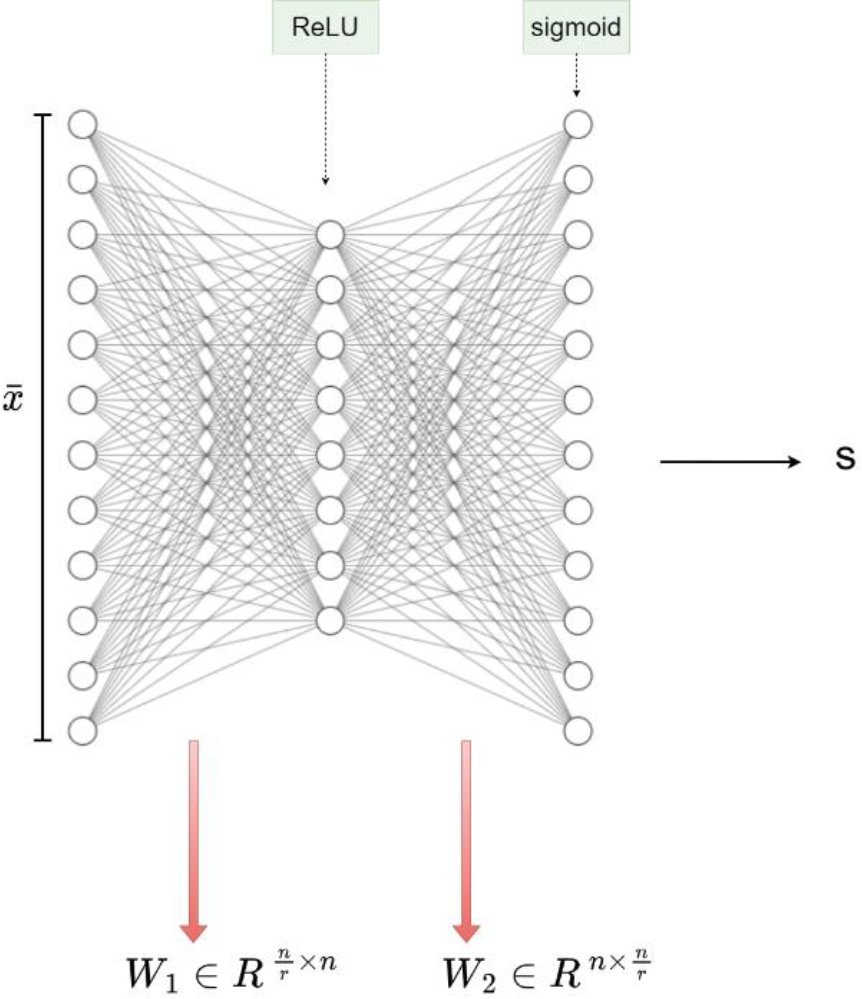
The excitation part of the SE block.

In SACNN, we also employed the residual block proposed in [39] with some modifications. Specifically, we replaced all the layers in the original residual block with their corresponding 1D counterparts, given that our inputs are unidimensional. Additionally, the second Conv1D layer in our residual architecture was assigned a dilation rate of 2 to capture a wider receptive field. The input tensor, which has been transformed by the Conv1D layers within the residual blocks, is further processed by a SE block. The SE block assigns greater influence on the more important channels of the input tensor, thereby enhancing the output. The resulting tensor is added to the original input signal, completing the residual connection within the block:

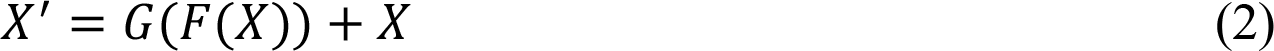

where *X* and *X*^′^ are the input and output of a residual block respectively, *F*(*X*) is the transformation function through the Conv1D layers in the residual block, and *G*(*X*) is the transformation function of the SE block.

Each standard Conv1D layer with a filter size of 3 captures the information of 3 consecutive points in each input. By stacking 2 of such layers, each output point can retrieve information from 5 consecutive points in the input. Conversely, a dilated Conv1D (dilation rate = 2) layer captures information from every other consecutive point (see Figure 5(b)). Thus, by stacking a standard Conv1D layer with a dilated Conv1D layer, each output point can access information from 7 points in the input layer. The difference between a standard Conv1D layer and a dilated Conv1D layer is illustrated in Figure 5.

**Figure 4.**
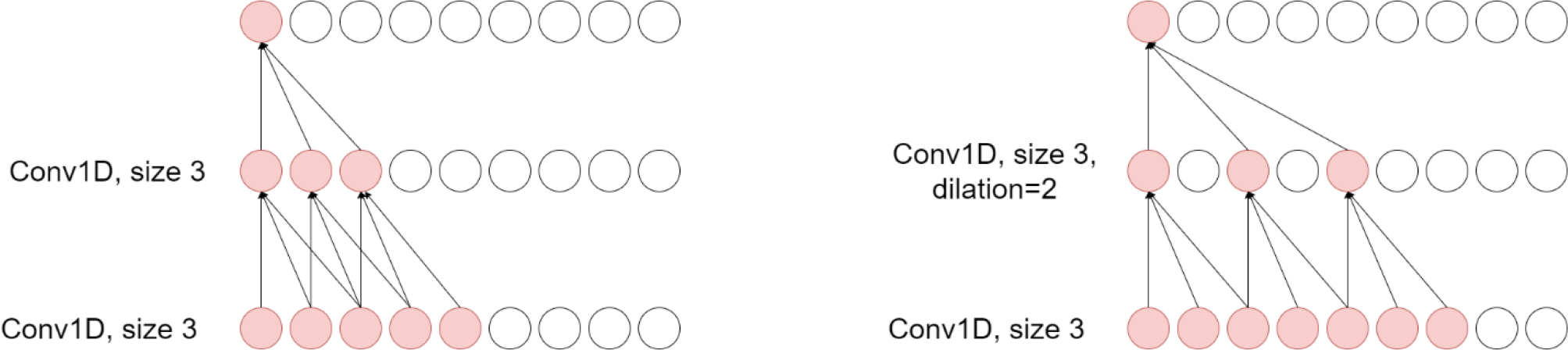
Comparison of a convolutional layer and dilated convolutional layer.

**Figure 5.**
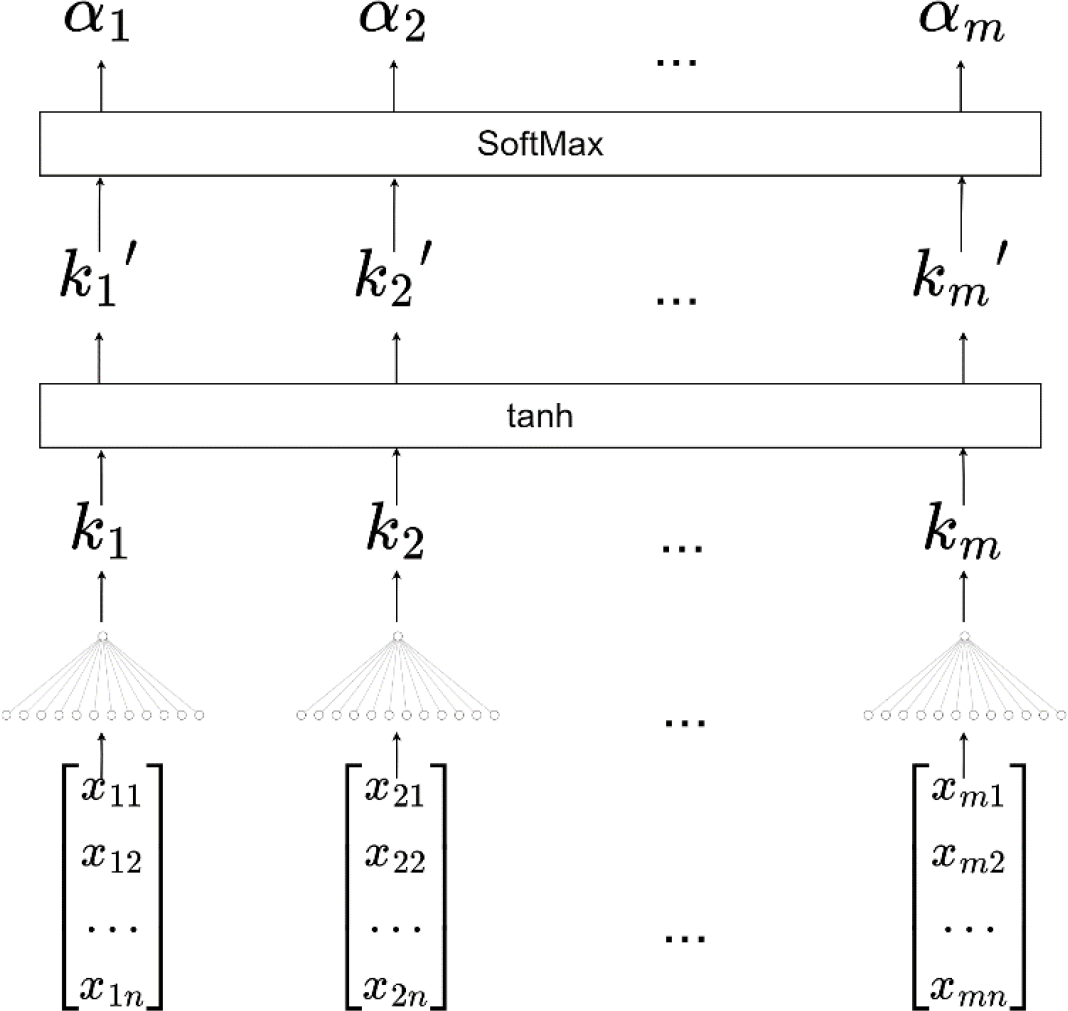
The process of generating the attention weights in the self-attention block in the SACNN

In this study, a single-head self-attention model was employed to analyze the extracted features through the SE-Res Blocks, with the aim of amplifying the impact of more important features on the final output. We use a method proposed in [40] to generate the attention weights. The equations below demonstrate how attention weights are calculated.

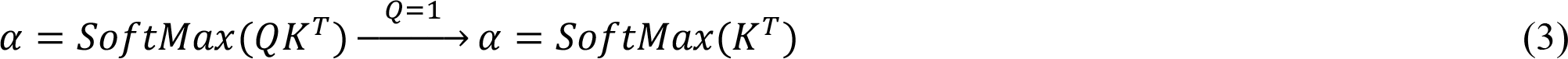

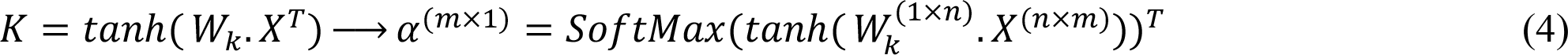

where α contains the attention weights, *X*^(*m*×*n*)^ is the input signal and to simplify the equation, *Q* is set to an adequate identical matrix. 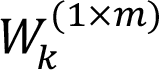 is defined during the training process by implementing a dense neural network. The SoftMax function ensures that the sum of attention weights is equal to 1. Figure 6 illustrates the process of generating attention weights in the attention block of our model.

**Figure 6.**
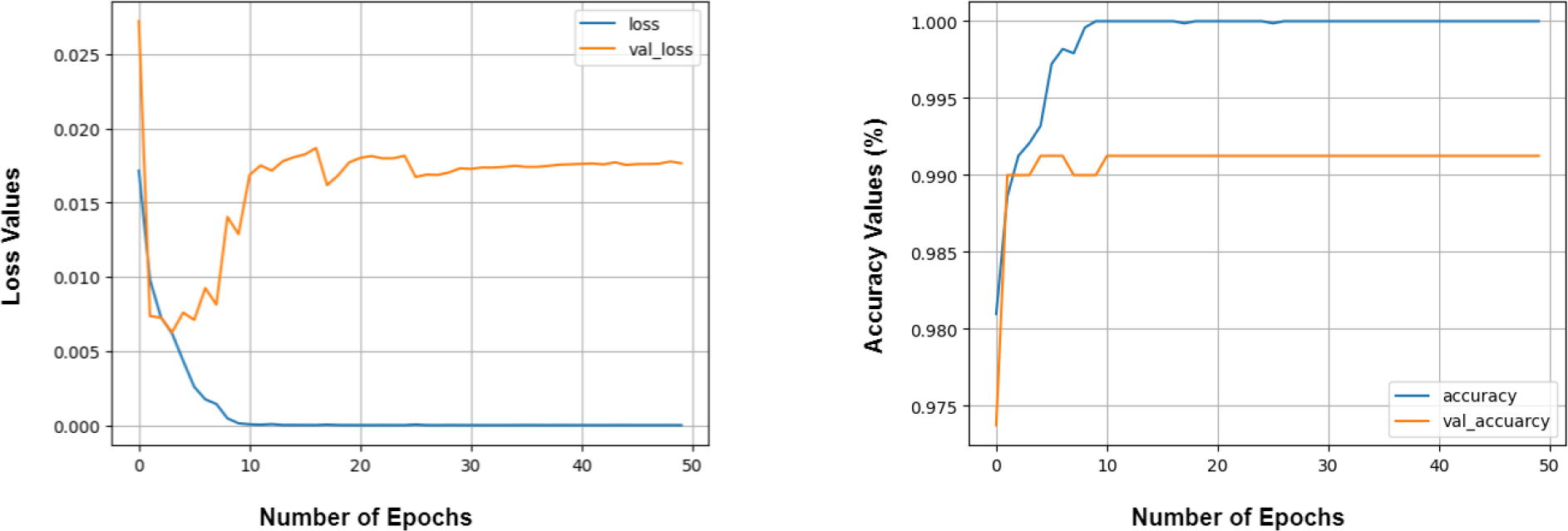
The results of SACNN on the training and the validation sets.

After calculating the attention weights, the output of the attention block, *X*^′^ ∈ *R*^*m*×*n*^, is generated by the equation below.

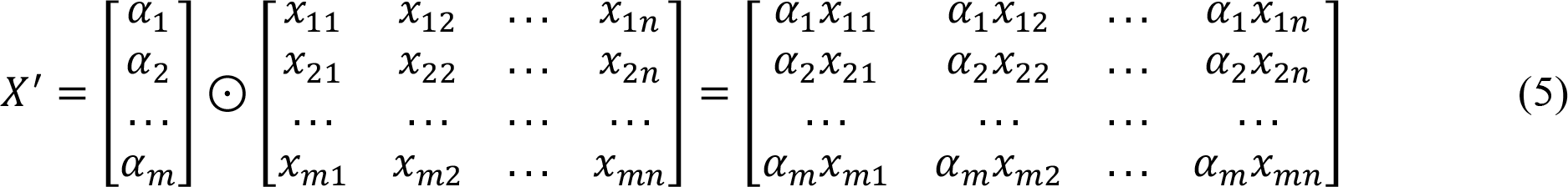

where ⊙ is element-wise multiplication.

It should be noted that each component in the output signal of the attention block in our model contains information from all the features of the input signal (via attention weights), which is not feasible using simple Conv1D or SE-Res blocks. This characteristic of the attention block enhances the performance of SACNN.

## 4. RESULTS

To evaluate the proposed model for diagnosing LBBB in 12-lead ECG signals, we used the dataset provided by the 2020 PhysioNet/Computing in Cardiology Challenge [35]. After preprocessing, the data was divided into a training-validation set consisting of 8,000 samples and a test set consisting of 2,344 samples. The data was split in such a way that both training-validation and test sets had an equal representation of LBBB. A stratified 10-fold cross-validation was applied to the training-validation set, with each fold using 90% of the data for training and the remaining 10% for validation. This stratified approach ensured that the proportion of LBBB samples was equal in the training and validation sets for all folds.

Due to the imbalanced data with only 2.23% of positive samples, it is crucial to choose appropriate metrics to evaluate the performance of the model. Therefore, we used multiple metrics, including accuracy, specificity, F1 score, and the area under the ROC curve, to assess the model’s performance [41]. The final classification of the model is determined by an optimal threshold, which was chosen by searching for a value that maximizes the F1 score, a more robust metric for imbalanced data. Ten different threshold values were obtained and evaluated for all ten models trained in different folds, resulting in a final optimal threshold of 0.488.

To establish a benchmark for comparison with the final results of the proposed model, several machine learning and deep learning models were implemented on the dataset. The machine learning models include Random Forest and Light Gradient Boosting Machine. These models require discrete features as inputs for classification. Therefore, statistical characteristics of two indicators from ECG signals (RR intervals and voltage values in R waves) are used as features. These statistical characteristics are first 5 percentile, first quartile, mean, third quartile, last 5 percentile, standard deviation, variance, and median. As a result, each sample was transformed into 192 discrete features (8 statistical characteristics of two indicators for 12 leads), which are used by the baseline models to classify a given sample. In addition to these machine learning models, deep learning models developed by Oh et al. [42] and Ribeiro et al. [28] are also applied to the dataset. They are popular deep learning models which perform properly on ECG signal classification task and can be considered as an appropriate benchmark. Table 1 presents the outcome of implementing the benchmark machine learning and deep learning models, which is used as a point of reference for evaluating the performance of proposed model. To ensure a fair comparison, all models were trained using a 10-fold cross-validation method. Furthermore, all deep models were trained for 50 epochs using Adam [43] (learning rate = 0.001) and Binary Cross Entropy as the optimizer and the loss function respectively. In all models, the final classification is performed by utilizing a threshold value of 0.5. Finally, we computed the same metrics for the SACNN’s on the test set. The results are tabulted in Table 1. As shown in Table 1, the SACNN outperforms the benchmarks in almost all metrics utilizing the same training hyperparameters. This observation strongly suggests that the architecture of SACNN is better suited for the task of 12-lead ECG classification.

**Table 1.**
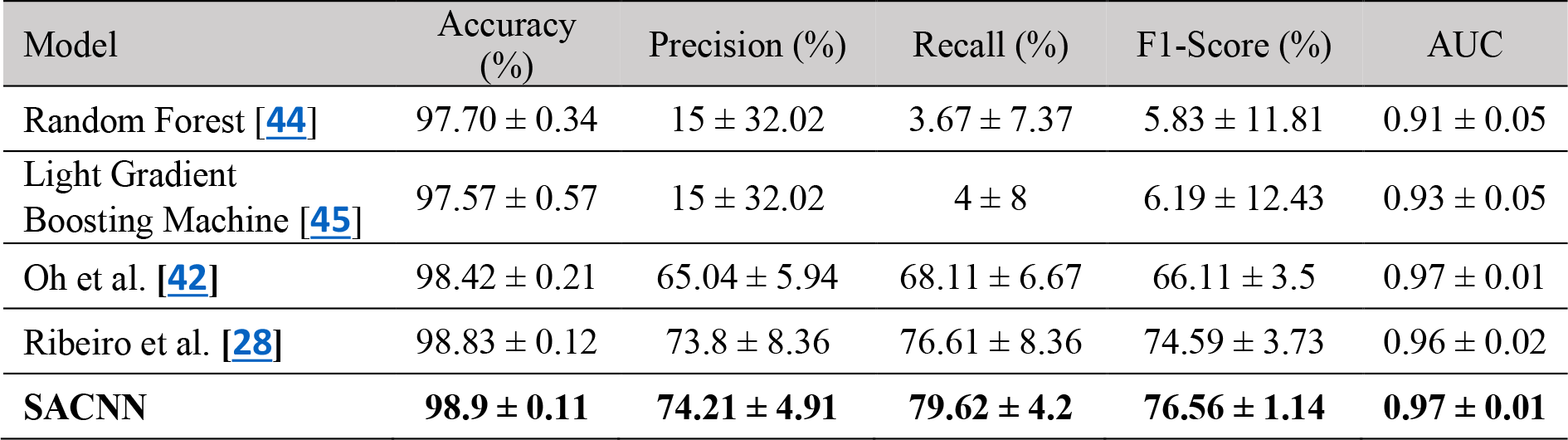
The performance of the benchmark models on detecting LBBB in the 12-lead ECG signals.

| Model | Accuracy (%) | Precision (%) | Recall (%) | F1-Score (%) | AUC |
| --- | --- | --- | --- | --- | --- |
| Random Forest [44] | $97.70 \pm 0.34$ | $15 \pm 32.02$ | $3.67 \pm 7.37$ | $5.83 \pm 11.81$ | $0.91 \pm 0.05$ |
| Light Gradient Boosting Machine [45] | $97.57 \pm 0.57$ | $15 \pm 32.02$ | $4 \pm 8$ | $6.19 \pm 12.43$ | $0.93 \pm 0.05$ |
| Oh et al. [42] | $98.42 \pm 0.21$ | $65.04 \pm 5.94$ | $68.11 \pm 6.67$ | $66.11 \pm 3.5$ | $0.97 \pm 0.01$ |
| Ribeiro et al. [28] | $98.83 \pm 0.12$ | $73.8 \pm 8.36$ | $76.61 \pm 8.36$ | $74.59 \pm 3.73$ | $0.96 \pm 0.02$ |
| **SACNN** | **$98.9 \pm 0.11$** | **$74.21 \pm 4.91$** | **$79.62 \pm 4.2$** | **$76.56 \pm 1.14$** | **$0.97 \pm 0.01$** |

To finetune SACNN, various optimizers, including Adam [43] (learning rate = 0.001), RMSProp [46] (learning rate = 0.001), and SGD (Learning rate = 0.001, momentum = 0.9), were tested and found that Adam performed the best, and was chosen as the optimizer. Moreover, during the training process, a learning rate scheduler was implemented, which began at a value of 10^-3^ for the first epoch and decreased uniformly concluding at a value of 10^-8^ for epoch number 50. The study also employed a focal binary cross-entropy loss function [47] to assess the degree of alignment between the predicted and actual classes of each sample. This loss function, which is a variant of the conventional binary cross-entropy, is particularly beneficial in classification tasks where the data is highly imbalanced. Additionally, the batch size and the number of epochs were set to the value of 32 and 50 respectively. The performance of SACNN after finetuning is as follows: accuracy of 98.91% ± 0.1%, specificity of 99.28% ± 0.15%, precision of 73.09% ± 3.5%, recall of 82.83% ± 4.34%, F1 score of 77.5% ± 1.59%, and AUC of 0.91 ± 0.02.

We also evaluated the learning curves of the SACNN on the dataset. Figure 7 illustrates the accuracy and loss curves for both the training and validation sets across the various epochs during training in Fold 1. The accuracy curve shows that the model achieved an impressive accuracy of 99% on the validation set in less than 5 epochs. Moreover, the loss curves indicate that the model was able to converge quickly and reach a satisfactory level of loss on the training and validation sets.

**Figure 7.**
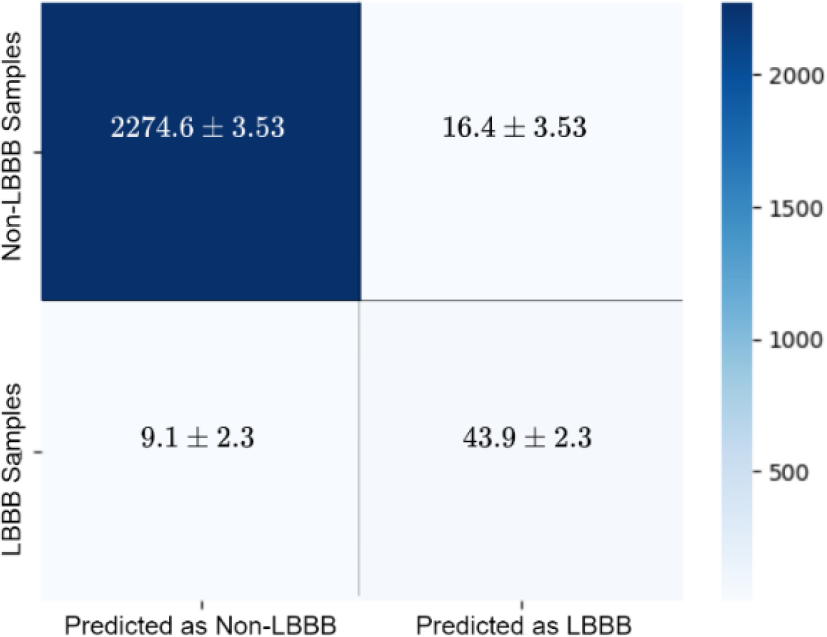
The confusion matrix of SACNN

The confusion matrices generated by the SACNN for diagnosing LBBB samples are presented in Figure 8. The number of false positives and false negatives is relatively low, with an average of only 16 false positives and 9 false negatives made by the SACNN.

**Figure 8.**
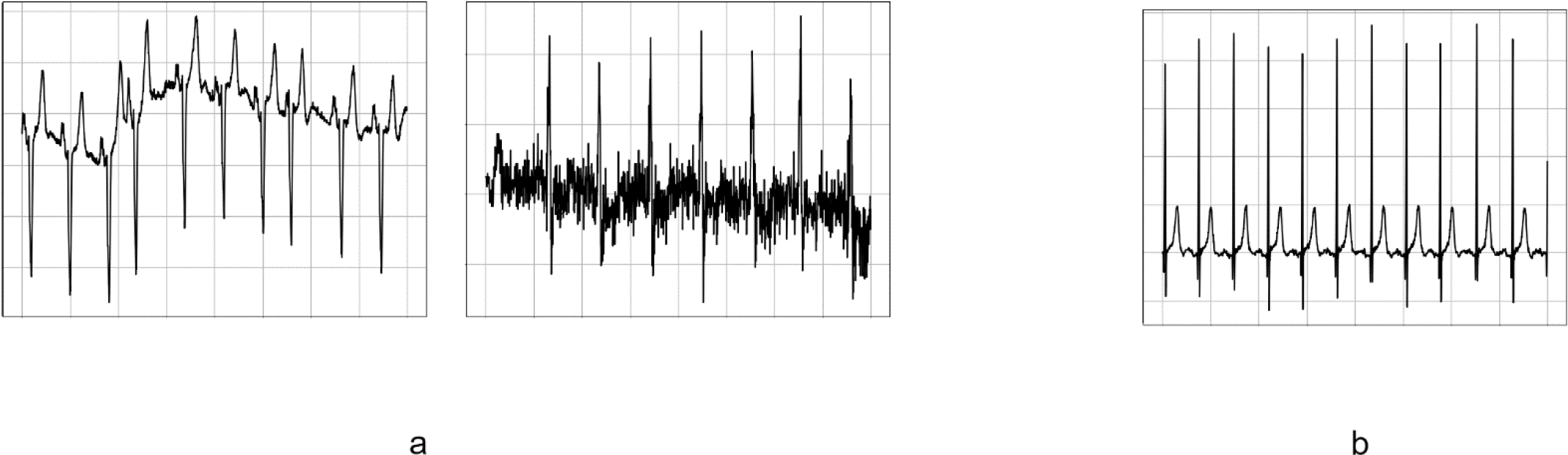
a) Lead aVF in 2 given samples misclassified in all 10 folds, b) Lead aVF in a random sample classified correctly.

Overall, the experimental results demonstrate that our deep learning model is effective for ECG signals on this dataset and holds great potential for other similar applications.

During the evaluation on the test set, it was found that 54 (2.1%) out of the 2,432 samples were misclassified at least once. Of these 54 misclassified samples, 13 (24.07%) were incorrectly classified in all 10 folds. To investigate the reasons of this misclassification, we took a look on these 13 samples. Figure 9 displays the lead V4 in two of these 13 samples that were misclassified in every fold and a randomly selected sample that was classified correctly.

After observing the two incorrectly classified samples and comparing them with a correctly classified sample, it was noted that there was some form of abnormality present in their ECG. This abnormality could be due to medical personnel’s mistake, patient noncompliance, or improper movements during ECG recording, or even due to the noise [48–50]. To further investigate this, the noise in the samples that were misclassified in all folds was removed using the Savitzky-Golay filter [51]. The model was then reapplied to the denoised samples. The results of this are shown in Table 2. Table 2 shows that the model’s ability to correctly classify all-fold-misclassified samples improves by over 20% through the denoising process. This shows that the model is more effective when there is less ECG defect present. However, we do not include the denoising to our training process as we aim to make the model robust to these effects and more prepared for the real-world circumstances.

**Table 2.** How all-fold-misclassified samples are classified befor and after denoising

|  |  | Sample 1 | Sample 2 | Sample 3 | Sample 4 | Sample 5 | Sample 6 | Sample 7 | Sample 8 | Sample 9 | Sample 10 | Sample 11 | Sample 12 | Sample 13 | Average |
| --- | --- | --- | --- | --- | --- | --- | --- | --- | --- | --- | --- | --- | --- | --- | --- |
| Number of Misclassified Folds | Before Denoising | 10 | 10 | 10 | 10 | 10 | 10 | 10 | 10 | 10 | 10 | 10 | 10 | 10 | **10** |
|  | After Denoising | 0 | 10 | 0 | 4 | 10 | 10 | 8 | 10 | 2 | 3 | 3 | 10 | 9 | **7.9** |

## 5. DISCUSSION

The proposed study presents a new deep learning model for accurately detecting LBBB, an arrhythmia in heart patients. The model is composed of SE-Res blocks and an attention block, and was validated using the 2020 PhysioNet/Computing in Cardiology Challenge data [35], which consists of 10,344 ECG samples with 2.23% suffering from LBBB. To ensure unbiased results, a ten-fold cross-validation method was used, and multiple performance metrics were applied to evaluate the performance of the system. The results showed that the proposed model achieved a high accuracy (98.91%) in detecting LBBB.

The proposed model was trained without using any noise-cancellation techniques, as this would have altered the structure of the ECG signals. This approach makes the model more practical for use in medical centers, as it can interpret the output of ECG recorders directly. In comparison with the benchmark machine learning models including logistic regression, random forest, and light gradient boosting machine, the proposed model outperforms them in all the evaluation metrics. The comparison proves that the proposed model is able to find the patterns in ECG signals which are more useful in diagnosing LBBB than the statistical characteristics of RR intervals that are commonly utilized by physicians.

A limitation of this study is that the patient’s clinical information, such as age, sex, and medical history, was not used in building the model. This information can play a role in the probability of a patient having LBBB arrhythmia, as the disease is more likely to occur in older individuals [52]. Unfortunately, such information was not fully provided in the used dataset.

## 6. CONCLUSION

This study introduces a novel deep neural network architecture called SACNN, which has been specifically developed for the purpose of detecting left bundle branch block (LBBB) from raw 12-lead electrocardiogram (ECG) recordings. Through comprehensive experimental evaluation, we have demonstrated the superior performance of SACNN in accurately identifying LBBB cases, surpassing other models designed for detecting heart arrhythmias. Furthermore, by evaluating the model’s performance on a large dataset, we have showcased the robust generalization capabilities of SACNN, thus highlighting its potential for real-world applications. It is worth noting that, in order to maintain the fidelity of the ECG inputs to actual raw recordings, fraud samples were included and no noise cancellation techniques were employed.

The findings of this research strongly support the suitability of the SACNN model for automatic and precise diagnosis of LBBB using 12-lead ECG data. This has significant implications for the future of LBBB screening and early detection in primary care settings, as it has the potential to alleviate the burden on healthcare professionals. Moreover, the SACNN architecture can be adapted and expanded to detect other cardiovascular conditions, thus broadening its impact on the field of cardiology and healthcare.

In conclusion, the SACNN model represents a significant advancement in the application of deep learning techniques for the detection of LBBB from 12-lead ECG recordings. This research not only sets the stage for more accurate and efficient LBBB diagnosis, but also serves as a foundation for future investigations into similar deep learning-based approaches for detecting and diagnosing other cardiovascular abnormalities.

## Data Availability

The study used ONLY openly available human data that were originally provided by the 2020 PhysioNet/Computing in Cardiology Challenge

